# Vitamin D and Central Blood Pressure: U-Shaped Risk and Copper Mediation

**DOI:** 10.1101/2025.10.13.25337949

**Authors:** Ching-Way Chen, Shu-Yu Tang, Yin-Yi Han, Sandy Huey-Jen Hsu, Jing-Shiang Hwang, Chi-Hua Cheng, Ta-Chen Su

**Author notes:** Correspondence Ta-Chen Su, MD, PhD, Professor and Deputy Superintendent, Tungs’ Taichung MetroHarbor Hospital, Address: No.699, Section 8, Taiwan Boulevard, Wuqi District,Taichung City 435403 Taiwan.

## Abstract

**Background:** Vitamin D deficiency has been implicated in cardiovascular risk, but its association with central blood pressure (cBP) and incident of hypertension in young Taiwanese adults remains unclear.

**Methods:** We analyzed 1,034 participants (mean age 33.45 years; 41.90% male) in the New YOTA cohort (2017–2019). Serum 25-hydroxyvitamin D [25(OH)D] was measured, with deficiency defined as <20 ng/mL. Associations of 25(OH)D with cBP were examined using multivariable regression.. Restricted cubic splines modeled the non-linear risk of hypertension. Structural equation modeling (SEM) assessed copper as a mediator.

**Results:** Vitamin D deficiency was observed in 54.1% of participants. Lower 25(OH)D was independently associated with higher central systolic, diastolic, and mean arterial pressure. A U-shaped with hypertension risk was observed, with the lowest risk at 20.1 ng/mL. Copper partially mediated the inverse relationship between 25(OH)D and cBP.

**Conclusion:** In young Taiwanese adults, vitamin D deficiency was linked to higher cBP and a U-shaped relationship with hypertension risk. Copper partially mediate this association. Maintaining adequate vitamin D levels may be important for supporting blood pressure regulation in early adulthood.

## Introduction

Hypertension is the third leading contributor to the global burden of disease^1^ and is projected to become increasingly prevalent with population aging and sedentary lifestyles. Elevated blood pressure (BP) is a major risk factor for cardiovascular disease (CVD), including coronary artery disease, myocardial infarction, and stroke and effective BP control remains essential for improving outcomes.^2^

Conventional BP assessment using a brachial cuff sphygmomanometer provides peripheral systolic and diastolic values, but captures only peak and trough pressures while omitting information from the entire arterial pressure waveform. By contrast, central blood pressure (cBP) is more pathophysiologically relevant to CVD development and hypertension-related organ damage.^3^

Vitamin D is an essential fat-soluble vitamin for its role in bone and mineral homeostasis,^4^ but its broader physiological effects have gained increasing attention. Serum 25-hydroxyvitamin D (25(OH)D) is the standard biomarker for assessing vitamin D status because of its relatively long half-life and stable serum concentration. According to the U.S. Institute of Medicine (2011) vitamin D is defined as 25(OH)D concentrations < 20 ng/mL (50 nmol/L).^5^ The Endocrine Society, in the same year, adopted the same threshold for deficiency but further categorized vitamin D insufficiency as 20–30 ng/mL (50–75 nmol/L) and sufficiency as > 30 ng/mL (75 nmol/L).^6^ The vitamin D receptor (VDR) is widely expressed in endothelial cells, vascular smooth muscle cells, renal tubular cells, nerve cells, dendritic cells and macrophages,^7^ implicating vitamin D in vascular biology, atherosclerosis, and regulation of the renin–angiotensin–aldosterone system (RAAS).^8^

Studies in vitamin D receptor (VDR) knockout mice demonstrate increased activity of the renin–angiotensin–aldosterone system (RAAS) and corresponding hypertension.^9^ In humans with normal blood pressure, lower vitamin D level have been associated with higher plasma renin activity, suggesting a role for vitamin D in RAAS regulation, although supplementation trials in hypertensive patients have not shown additive benefits when combined with standard antihypertensive therapy.^10,11^

Epidemiologic studies provide further evidence linking vitamin D to blood pressure regulation. In U.S. cohorts, lower vitamin D concentrations were inversely associated with systolic blood pressure (SBP), though this relationship was attenuated after age adjustment.^12,13^ The National Health and Nutrition Examination Survey (NHANES) III study demonstrated a stronger inverse association in adults over 50 years.^14^ Similar findings have been reported in Germany,^15^ Korea, and pediatric populations, where lower vitamin D levels correlated with higher SBP or diastolic blood pressure (DBP), often influenced by adiposity.^16^ Prospective studies suggest that higher vitamin D levels are associated with reduced risk of incident hypertension,^17^ whereas lower levels predict future hypertension, as shown in middle-aged Turkish adults.^18^

Copper, an essential micronutrient and enzymatic cofactor, may also play a role in hypertension.^19^ While adequate copper is necessary for physiological function, excessive exposure has been linked to oxidative stress, endothelial dysfunction, and increased blood pressure. Both animal and human studies have suggested that elevated serum copper concentration are linked to hypertension.^20^ Moreover, dietary copper intake has been associated with blood pressure regulation, with evidence showing a U-shaped relationship between intake and new-onset hypertension.^21^ In addition, Mendelian randomization analyses and data from the NHANES have suggested that higher copper intake is associated with an increased risk of developing hypertension.^22^

Our previous research indicated that copper may mediate the association between vitamin D deficiency and impaired pulmonary function.^23^ However, whether copper mediates the relationship between vitamin D and hypertension remains unclear. In Taiwan, vitamin D deficiency affects approximately 22% of general population.^25^ Long-term cohort data have shown that lower serum 25-hydroxyvitamin D [25(OH)D] predicts both cardiovascular and all-cause mortality among individuals with diabetes.^24,25^ Despite these findings, no study has specifically examined the association between vitamin D status and cBP, a measure more closely related to target organ damage, in young, healthy Taiwanese adults. Therefore, we investigated the relationship between vitamin D status and cBP in this population and further explored whether copper serves as a mediator in this association.

## Material and Methods

### Study Population

Between 1992 and 2000, a national urine screening program in Taiwan enrolled approximately 2.6 to 2.9 million school-aged children (6–18 years). Of these, 103,756 participants underwent follow-up, including 9,227 with elevated blood pressure and 94,529 with normal levels.^26^ From this program, the Young Taiwanese Cohort (YOTA) as established in 2006–2008and has been described previously.^27,28^

From 2017 to 2019, we recontacted YOTA participants and recruited additional volunteers from the general population in Taipei, yielding a total of 1,034 adults (542 from the original YOTA and 492 community volunteers). The study was approved by the Institutional Review Board of National Taiwan University Hospital (IRB No. 201604089RINA), and written informed consent was obtained from all participants. Individuals with active cancer, cognitive impairment, or who declined participation were excluded.^28^

### Assessment of clinical information and risk stratification

Participants underwent standardized evaluation of cardiovascular risk factors. Age, gender, and anthropometric measurements were recorded, and body mass index (BMI) was calculated as weight (kg)/height (m²). Fasting blood and urine samples were obtained to measure serum creatinine, glycated hemoglobin (HbA1c), insulin resistance (HOMA-IR), low-density lipoprotein cholesterol (LDL-C), and total cholesterol (TCHO)..

Lifestyle information was collected using self-administered questionnaires. Smoking was defined as regular use of tobacco, alcohol consumption as ≥2 drinks per week, and regular exercise as ≥90 minutes of physical activity per week.

### Cardiac and vascular function assessments

We used a cuff sphygmomanometer equipped with an oscillometric device (DynaPulse 200M, Pulse Metric Inc., San Diego, CA, USA) to measure the arterial pressure waveform.^29^ Measurements were obtained from the left and right arms after at least 5 minutes of rest in a sitting position in a quiet classroom. The average of two measurements was used in the analyses. The Pulse-Dynamics technology analyzes the oscillometric brachial arterial pressure waveforms acquired from the cuff and, by pulse-waveform analysis, determines central systolic blood pressure (cSBP), central diastolic blood pressure (cDBP), and central mean arterial pressure (cMAP).^30–32^ The device records arterial pressure waveforms from a standard brachial cuff and analyzes waveform changes using Bernoulli’s principle. This method has been validated against invasive measurements and shown to provide accurate estimates of central hemodynamics and arterial stiffness.^33^ It has also been employed in our group’s previous studies.^29,34,35^ Hypertension was defined according to the 2022 Taiwan Hypertension Guidelines^36^ and 2025 AHA/ACC Guideline for the Prevention, Detection, Evaluation, and Management of High Blood Pressure in Adults^37^ as an average BP ≥130/80 mmHg.

### Quantification of serum vitamin D levels

Venous blood samples were collected after an overnight fast of approximately 12 hours and stored at −80℃ until analysis. Serum 25(OH)D concentration, calculated as the sum of 25(OH)D₂ and 25(OH)D₃, was measured using the TOTAL Liaison chemiluminescent assay (Liaison, Diasorin S.p.A., Saluggia, Italy).^38^ The assay demonstrated a coefficient of variation of 2.65% (SD = 1.2%).^39^ To minimize seasonal variation, recruitment and sample collection were conducted between March– September of each study year (2017–2019).

Urinary copper concentrations were measured using inductively coupled plasma mass spectrometry (ICP-MS; 7700 series, Agilent Technologies, Santa Clara, CA, USA) according to established protocols. For samples below the detection limit, values were imputed as one-half of the limit of detection (0.276 μg/L). All analyses were performed in an accredited laboratory (National Environmental Laboratory Accreditation Conference, NELAC 2003) and certified through the international quality assurance program (G-EQUAS-67).

### Statistical analysis

The minimum sample size was estimated at 385 participants, assuming a 50% prevalence of vitamin D deficiency (25[OH]D <20 ng/mL), 95% confidence level, and 5% margin of error. Data normality was assessed using the Shapiro–Wilk and Kolmogorov–Smirnov tests.. Normally distributed variables were reported as mean ± SD, and comparisons between groups were performed using Student’s *t* test for continuous variables and χ² test for categorical variables.

Associations between serum 25(OH)D and cBP were examined using linear regression. Variables with *P* < 0.10 in univariate analyses were considered in multivariable modeling. Four nested models were constructed: Model 1 unadjusted; Model 2 adjusted for age and gender; Model 3 further adjusted for body mass index, serum creatinine, HbA1c, LDL-C, total cholesterol, and albumin levels; and Model 4 additionally adjusted for smoking and exercise. Subgroup analyses stratified by age (≥30 vs. <30-years), gender, BMI (≥24 vs. <24 kg/m²), smoking status, and regular exercise were conducted. Interaction terms were tested to assess effect modification. Non-linear associations between 25(OH)D and hypertension risk were evaluated using restricted cubic spline (RCS) logistic regression models with the rcs() function in R. Analyses were performed in the overall cohort and stratified by gender and age groups (women <30 years, men <30 years, women ≥30 years, and men ≥30 years). Non-linearity and interaction effects were tested using Wald statistics. Given the low prevalence of hypertension, parsimonious models were applied (unadjusted and adjusted for age and gender). Finally, mediation analysis was conducted using structural equation modeling (SEM) to evaluate the role of urinary copper in the association between 25(OH)D and cBP. Analyses were performed using SPSS (version 28.0; IBM, Armonk, NY, USA) and R (version 4.3.1; R Core Team, 2024). A two-tailed *P*<0.05 was considered statistically significant.

## Results

Our study included 1,034 participants (mean age, 33. 5 ±7.9 years), of whom 428 (41.4%) were men and 606 (58.6%) were women. The median serum 25(OH)D concentration was 19.2 ng/mL (IQR, 11.5–24.9 ng/mL). Based on both the cohort median and established guideline thresholds, participants were classified into two groups using a cut-off of 20 ng/mL.

Compared with those with 25(OH)D ≥20 ng/mL, participants with 25(OH)D <20 ng/mL were younger (32.5 ± 6.6 vs 34.8 ± 9.1 years, *P* < 0.01) and less likely to be men (34.4% vs 49.7%, *P* < 0.01). They also had higher LDL-C level (109.0 ± 33.1 vs 104.8 ± 30.2 mg/dL, *P* = 0.03) and higher total cholesterol concentration (192. 3 ± 46.7 vs 184.7 ± 34.1 mg/dL, *P* < 0.01). In contrast, participants with 25(OH)D ≥ 20 ng/mL were more likely to be smokers (16.0% vs 11. 5%, *P* = 0.03) and to report regular exercise (63.4% vs 55.6%, *P* < 0.01). Detailed baseline characteristics are shown in Table 1.

**Table 1.** Characteristics of vitamin D deficient and non-deficient groups.

| Variables | Total Population<br>N=1034 | Vitamin D Deficiency<br>25(OH)D < 20 ng/mL<br>N=559 (54.06%) | Vitamin D Non-Deficient<br>25(OH)D ≥ 20 ng/mL<br>N=475 (45.94%) | *P* value |
| --- | --- | --- | --- | --- |
| Age (years) | 33.45 ± 7.91 | 32.31 ± 6.56 | 34.83 ± 9.10 | < 0.01 |
| Men (%) | 428 (41.39%) | 192 (34.35%) | 236 (49.68%) | < 0.01 |
| BMI (kg/m<sup>2</sup>) | 23.08 ± 4.23 | 23.04 ± 4.35 | 23.14 ± 4.08 | 0.70 |
| cSBP (mmHg) | 114.06 ± 15.01 | 114.87 ± 14.23 | 113.09 ± 15.64 | 0.05 |
| cDBP (mmHg) | 65.21 ± 9.82 | 65.75 ± 9.65 | 64.55 ± 9.98 | 0.05 |
| HTN (%) | 41 (3.96%) | 18 (3.22%) | 23 (4.84%) | 0.15 |
| Creatinine (mg/dL) | 0.88 ± 0.92 | 0.91 ± 1.23 | 0.86 ± 0.21 | 0.40 |
| Glucose (mg/dL) | 87.38 ± 20.41 | 87.75 ± 25.16 | 86.92 ± 12.48 | 0.52 |
| HbA1C (%) | 5.66 ± 0.70 | 5.65 ± 0.81 | 5.67 ± 0.55 | 0.61 |
| LDL-C (mg/dL) | 107.09 ± 31.85 | 109.01 ± 33.05 | 104.77 ± 30.21 | 0.03 |
| TCHO (mg/dL) | 188.87 ± 41.61 | 192.33 ± 46.66 | 184.68 ± 34.12 | < 0.01 |
| Albumin (g/dL) | 4.71 ± 0.27 | 4.70 ± 0.28 | 4.72 ± 0.25 | 0.32 |
| Calcium (mg/dL) | 9.33 ± 1.31 | 9.29 ± 0.43 | 9.39 ± 1.89 | 0.24 |
| Phosphate (mg/dL) | 3.65 ± 3.03 | 3.60 ± 0.61 | 3.72 ± 4.45 | 0.52 |
| Smoking (%) | 140 (13.54%) | 64 (11.45%) | 76 (16.00%) | 0.03 |
| Alcohol (%) | 321 (31.50%) | 174 (31.13%) | 147 (30.95%) | 0.87 |
| Exercise (%) | 612 (59.30%) | 311 (55.64%) | 301 (63.37%) | < 0.01 |
| Education (%) | 910 (88.01%) | 504 (90.16%) | 406 (85.47%) | 0.30 |
| Supplement* (%) | 133 (12.86%) | 66 (11.81%) | 67 (14.11%) | 0.24 |

Abbreviation: 25(OH)D: 25-hydroxyvitamin D; BMI: body mass index; cSBP: central systolic blood pressure; cDBP: central diastolic blood pressure; HOMA-IR: homeostatic model assessment got Insulin resistance; hs-CRP: high sensitivity C- reactive protein; LDL-C: low-density lipoprotein cholesterol; TCHO: total cholesterol; Smoking: active smoker (Yes/ No); Exercise: regular exercise habit (Yes/No); Education: Bachelor's degree or higher
Data presented as mean ± standard deviation for continuous variables and N (%) for categorical variables.
\* Oral calcium supplement

### Univariate Analysis

As shown in Table 2, lower serum 25(OH)D concentrations were significantly associated with higher cSBP and cDBP. Higher cSBP was also associated with male gender, higher BMI, elevated serum creatinine, fasting glucose, HbA1C, LDL-C, smoking, and higher educational attainment. Regular exercise showed a borderline association with lower cSBP (*P* = 0.05), whereas age was inversely associated with cSBP (*P* < 0.01). For cDBP, similar associations were observed, with the exception that total cholesterol was significant (*P* < 0.01), whereas serum albumin and education were not. Alcohol consumption was not significantly associated with either cSBP or cDBP (Table 2).

**Table 2.** The relationship between central systolic blood pressure (cSBP) and central diastolic blood pressure (cDBP) and cardiovascular risk factors using univariate linear regression.

|  | cSBP (mmHg) |  |  | cDBP (mmHg) |  |  |
| --- | --- | --- | --- | --- | --- | --- |
|  | Beta | SE | *P* value | Beta | SE | *P* value |
| 25(OH)D (ng/mL) | −0.08 | 0.05 | 0.01 | −0.13 | 0.03 | < 0.01 |
| Age (years) | −0.54 | 0.06 | < 0.01 | −0.11 | 0.04 | < 0.01 |
| Men (%) | 12.43 | 0.87 | < 0.01 | 6.15 | 0.59 | < 0.01 |
| BMI (kg/m<sup>2</sup>) | 1.70 | 0.10 | < 0.01 | 1.04 | 0.07 | < 0.01 |
| Creatinine (mg/dL) | 2.39 | 0.50 | < 0.01 | 1.41 | 0.33 | < 0.01 |
| Glucose (mg/dL) | 0.08 | 0.02 | < 0.01 | 0.06 | 0.02 | < 0.01 |
| HbA<sub>1c</sub> (%) | 1.45 | 0.66 | 0.03 | 1.32 | 0.43 | < 0.01 |
| LDL-C (mg/dL) | 0.08 | 0.01 | < 0.01 | 0.07 | 0.01 | < 0.01 |
| TCHO (mg/dL) | 0.02 | 0.01 | 0.123 | 0.02 | 0.01 | < 0.01 |
| Albumin (g/dL) | 5.81 | 1.75 | < 0.01 | 1.78 | 1.15 | 0.12 |
| Smoking (%) | 4.91 | 1.36 | < 0.01 | 3.19 | 0.89 | < 0.01 |
| Alcohol (%) | 0.44 | 1.01 | 0.66 | 0.75 | 0.66 | 0.26 |
| Exercise (%) | −1.88 | 0.95 | 0.05 | −1.23 | 0.62 | 0.05 |
| Education (%) | 4.43 | 1.43 | < 0.01 | 0.82 | 0.94 | 0.39 |

Abbreviation: 25(OH)D: 25-hydroxyvitamin D; cSBP: central systolic blood pressure; cDBP: central diastolic blood pressure; BMI: body mass index; HbA<sub>1c</sub>: glycated hemoglobin; LDL-C: low-density lipoprotein cholesterol; TCHO: total cholesterol; Smoking: active smoking (yes/no); Alcohol: regular social drinking (yes/no); Exercise: regular exercise (yes/no); Education: Bachelor's degree or higher (yes/no); Beta: slope parameter; SE: standard error

### Multivariable Analysis

In multivariable linear regression, serum 25(OH)D remained inversely associated with cSBP, cDBP, and cMAP across all four models. In the fully adjusted model (Model 4), each 10 ng/mL increase in 25(OH)D was associated with a −0.8 mmHg change in cSBP (standard error [SE] = 0.4, *P* = 0.04), −0.9 mmHg change in cDBP (SE = 0.3, *P* = 0.02), and −1.0 mmHg change in cMAP (SE = 0.3, *P* = 0.03) (Table 3).

**Table 3.** Multivariable linear regression analysis of the association between central systolic blood pressure (cSBP), central diastolic blood pressure (cDBP), and central mean arterial pressure (cMAP) and serum 25(OH)D concentrations.

|  | cSBP (mmHg) |  |  | cDBP (mmHg) |  |  | cMAP (mmHg) |  |  |
| --- | --- | --- | --- | --- | --- | --- | --- | --- | --- |
|  | Beta | SE | *P* value | Beta | SE | *P* value | Beta | SE | *P* value |
| Model 1 | −0.08 | 0.05 | 0.01 | −0.13 | 0.03 | < 0.01 | −0.09 | 0.04 | < 0.01 |
| Model 2 | −0.08 | 0.05 | 0.02 | −0.08 | 0.03 | < 0.01 | −0.15 | 0.04 | < 0.01 |
| Model 3 | −0.09 | 0.04 | 0.03 | −0.09 | 0.03 | 0.01 | −0.10 | 0.03 | 0.02 |
| Model 4 | −0.08 | 0.04 | 0.04 | −0.09 | 0.03 | 0.02 | −0.10 | 0.03 | 0.03 |

Abbreviation: cSBP: central systolic blood pressure; cDBP: central diastolic blood pressure; cMAP: central mean arterial pressure; Beta: slope parameter; SE: standard error
Model 1: Unadjusted; Model 2: Adjusted for age and gender; Model 3: Adjusted for age, gender, body mass index, serum creatine level, glycated hemoglobin (HbA1C), low-density lipoprotein cholesterol, total cholesterol, serum albumin concentration; Model 4: Adjusted for age, gender, BMI, serum creatinine level, HbA1C, LDL-C, TCHO, serum albumin concentration, cigarette smoking, and regular exercise.

### Subgroup Analysis

The inverse association between 25(OH)D and cSBP was more pronounced in women and in participants with BMI < 24 kg/m², and was also stronger among those reporting regular exercise. A similar trend was observed in non-smokers, although it did not reach statistical significance. Association between 25(OH)D and cBP were consistent across age strata (< 30 vs. ≥ 30 years) (Table 4).

**Table 4.** Subgroup analysis detailing the relationship between 25(OH)D concentrations and the central systolic blood pressure (cSBP) using multivariable linear regression.

| Subgroup | Number | cSBP (mmHg) |  |  |  |
| --- | --- | --- | --- | --- | --- |
|  |  | Beta | SE | *P* value | *P* for interaction |
| Overall | 1034 | −0.08 | 0.04 | 0.04 |  |
| Gender |  |  |  |  | < 0.01* |
| Men | 428 | −0.07 | 0.06 | 0.27 |  |
| Women | 606 | −0.10 | 0.06 | 0.08* |  |
| Age |  |  |  |  | 0.34 |
| ≥ 30 y/o | 663 | −0.19 | 0.06 | < 0.01* |  |
| < 30 y/o | 371 | −0.10 | 0.06 | 0.08* |  |
| BMI |  |  |  |  | < 0.01* |
| ≥ 24 kg/m<sup>2</sup> | 370 | −0.21 | 0.09 | 0.80 |  |
| < 24 kg/m<sup>2</sup> | 664 | −0.10 | 0.05 | 0.03* |  |
| Smoking |  |  |  |  | 0.79 |
| Yes | 140 | −0.13 | 0.12 | 0.30 |  |
| No | 894 | −0.08 | 0.04 | 0.08* |  |
| Exercise |  |  |  |  | < 0.01* |
| Yes | 613 | −0.12 | 0.06 | 0.03* |  |
| No | 421 | −0.03 | 0.06 | 0.58 |  |

Abbreviation: cSBP: central systolic blood pressure; BMI: body mass index; Beta: slope parameter; SE: standard error
Multivariable linear regression: Adjusted for age, gender, BMI, serum creatinine level, HbA<sub>1c</sub>, LDL-C, TCHO, serum albumin concentration, cigarette smoking, and regular exercise. The specific variable was not included as a covariate in its respective subgroup analysis.
*P* for interaction: The *P* value for interaction assesses whether the effect of the independent variable (25(OH)D concentration) on the outcome (systolic blood pressure) differs significantly between subgroups (e.g., gender, age, BMI, smoking, regular exercise).
\* *P* value < 0.05

### Restricted Cubic Spline Analysis

Restricted cubic spline regression demonstrated a significant non-linear association between serum 25(OH)D and hypertension risk. (P for non-linearity= 0.01; P for linearity=0.60). The lowest predicted risk was observed at 20.1 ng/mL, corresponding to a 12.3% risk of hypertension (95% CI, 10.4–14.6%) (Figure 1A). The U-shaped pattern suggested that both lower and higher 25(OH)D concentration were associated with increased risk of developing hypertension. After adjustment for age and gender, the nadir of risk shifted to 23.6 ng/mL, where the odds ratio of hypertension was 0.86 (95% CI, 0.72–1.03), relative to a reference at 20.1 ng/mL (Figure 1B).

**Figure 1.**
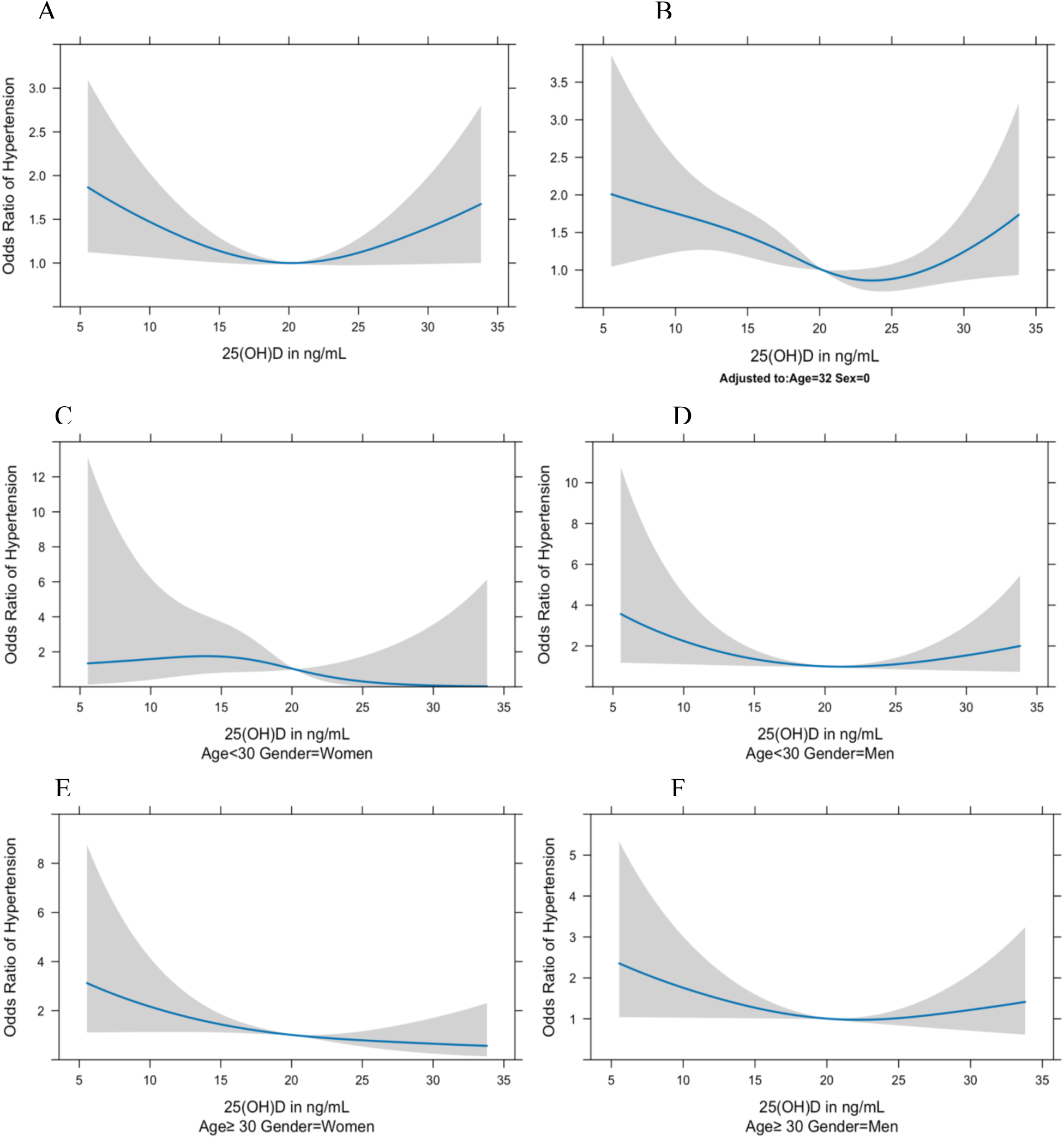
Non-linear associations between serum 25(OH)D concentrations and hypertension risk in the overall sample and in age–gender subgroups. Restricted cubic spline (RCS) regression models showing odds ratios (ORs) for hypertension across serum 25-hydroxyvitamin D [25(OH)D] concentrations in the overall sample and by age–gender subgroups. Panels A and B display the overall association without and with adjustment for age and gender, respectively. In Panel A, the non-linear association was significant (*P* = 0.01; *P* for linearity = 0.60), with the lowest predicted risk at 20.1 ng/mL (12.31%, 95% CI: 10.4–14.6%). In Panel B, after adjusting, the the non-linear association remained significant (*P* < 0.01), with minimal risk at 23.6 ng/mL (OR = 0.86, 95% CI: 0.72–1.03) relative to 20.1 ng/mL. Panels C–F show subgroup-specific associations stratified by age and sex. Signification non-linear associations were observed in men <30 years (Panel D, *P* = 0.02) and ≥30 years (Panel F, *P* = 0.03), but not in women (Panel C and Panel E). Despite these patterns, formal interaction tests between vitamin D, age, and gender were not statistically significant (See Table 5). The Y-axis represents ORs of hypertension; the X-axis shows serum 25(OH)D concentrations ng/mL. Values > 1 indicate increased risk, values < 1 indicate reduced risk, and shaded areas represent 95% confidence intervals.

**Table 5.**
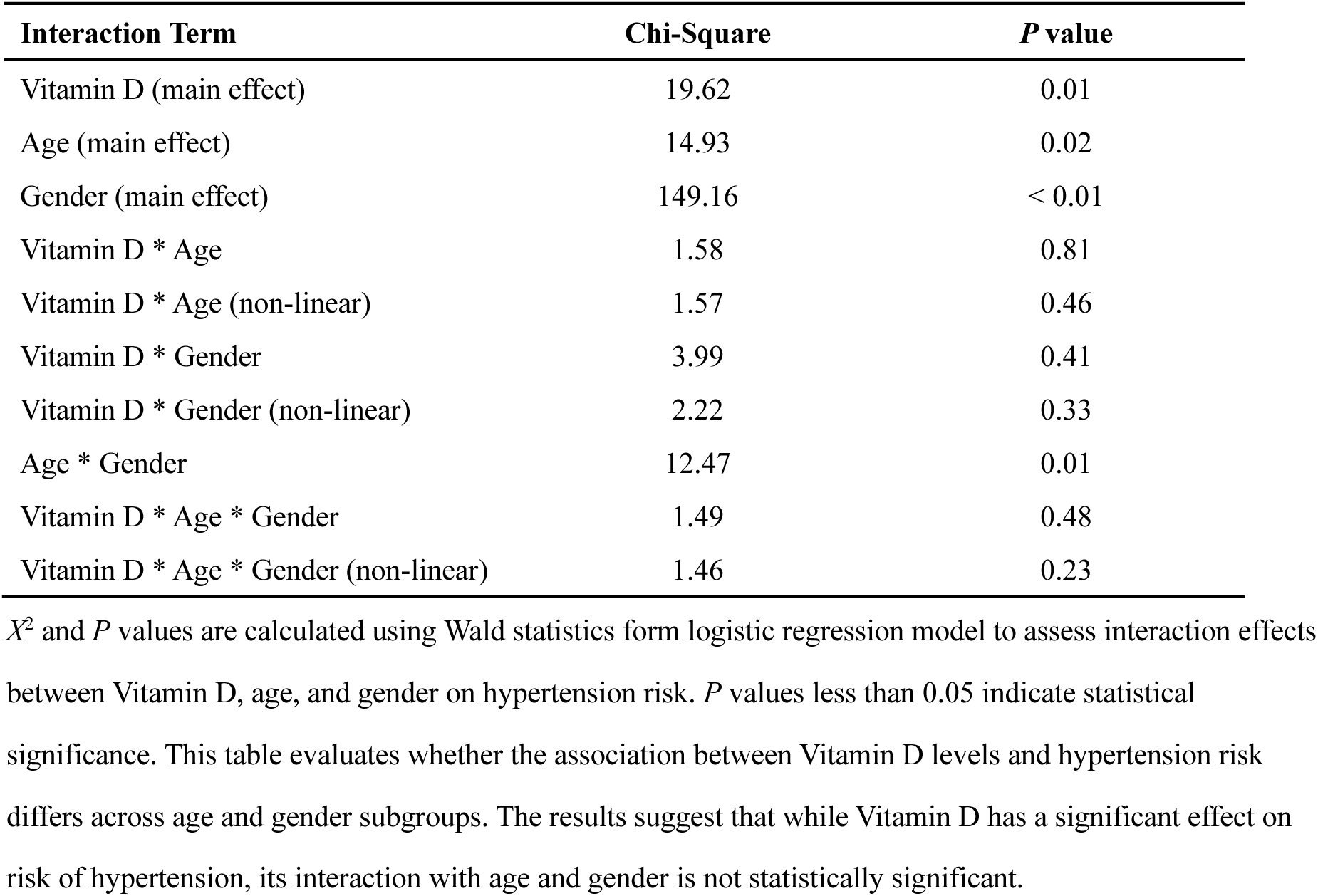
Interaction effects of vitamin D, age, and gender on risk of hypertension (refer to **Figure 1**)

| **Interaction Term** | **Chi-Square** | ***P* value** |
| --- | --- | --- |
| Vitamin D (main effect) | 19.62 | 0.01 |
| Age (main effect) | 14.93 | 0.02 |
| Gender (main effect) | 149.16 | < 0.01 |
| Vitamin D * Age | 1.58 | 0.81 |
| Vitamin D * Age (non-linear) | 1.57 | 0.46 |
| Vitamin D * Gender | 3.99 | 0.41 |
| Vitamin D * Gender (non-linear) | 2.22 | 0.33 |
| Age * Gender | 12.47 | 0.01 |
| Vitamin D * Age * Gender | 1.49 | 0.48 |
| Vitamin D * Age * Gender (non-linear) | 1.46 | 0.23 |

$\chi^2$ and *P* values are calculated using Wald statistics from logistic regression model to assess interaction effects between Vitamin D, age, and gender on hypertension risk. *P* values less than 0.05 indicate statistical significance. This table evaluates whether the association between Vitamin D levels and hypertension risk differs across age and gender subgroups. The results suggest that while Vitamin D has a significant effect on risk of hypertension, its interaction with age and gender is not statistically significant.

Stratified analyses revealed significant non-linear associations among men < 30 years (p=0.02) and ≧30 years (p=0.03), with the lowest predicted risk at 24.0 ng/mL and 23.7 ng/mL, respectively (Figure 1D, 1F). In contrast, no significant non-linear association were observed among women, wither <30 years (p=0.03) or ≥ 30 years (p=0.64) (Figure 1C, 1E). Although the risk patterns appeared stronger in men, formal interaction testing indicated no significant modification by age or gender.

### Interaction Analysis

As shown in Table 5, main effects of 25(OH)D (χ² = 19.62, *P* = 0.01), age (χ² = 14.93, *P* = 0.02), and gender (χ² = 149.16, *P* < 0.01) were significant, indicating independent contributions to hypertension risk. However, no significant interactions were observed between 25(OH)D and age (χ² = 1.58, *P* = 0.81) or gender (χ² = 3.99, *P* = 0.41) nor for their non-linear terms (*P* = 0.46 and *P* = 0.33). The age-gender interaction was significant (χ² = 12.47, *P* = 0.01), but the three-way interaction with 25(OH)D was not (*P* = 0.48; non-linear *P* = 0.23) (Table 5).

### Structural equation modeling (SEM)

SEM analyses showed that serum 25(OH)D was inversely associated with urinary copper, which in turn was positively associated with cSBP. For cSBP, the direct effect of 25(OH)D was −0.076, the indirect effect via copper was −0.004, and the total effect was −0.080 (Table S1). Model fit was acceptable (GFI = 0.93, NFI= 0.95, RMSEA= 0.07; Table S3), and the SEM model is illustrated in Figure 2.

**Figure 2.**
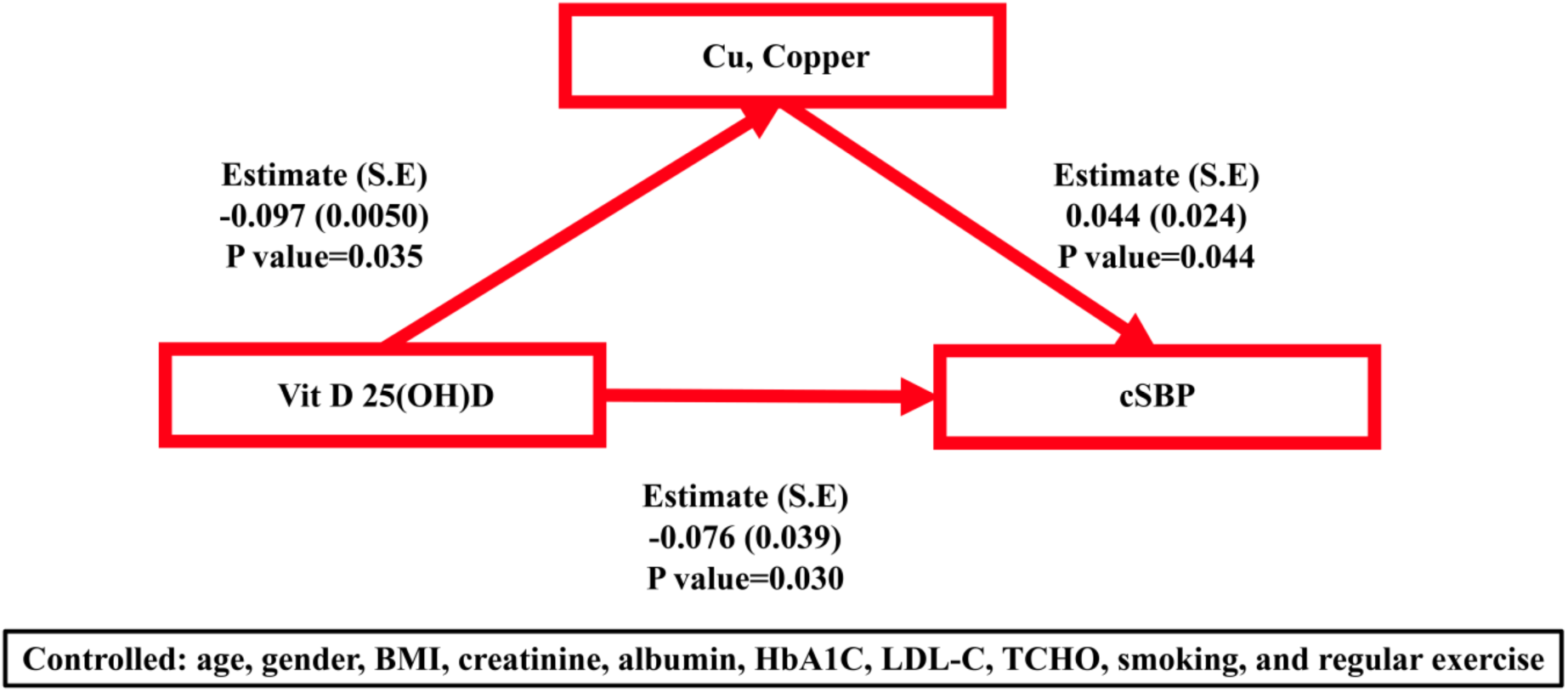
Structural equation modeling (SEM) illustrating the associations among serum 25-hydroxyvitamin D [25(OH)D], urinary copper (Cu), and central systolic blood pressure (cSBP). Arrows indicate direct and indirect pathways with corresponding standardized coefficients (standard errors) and P values. The model was adjusted for age, gender, body mass index, serum creatinine, albumin, glycated hemoglobin (HbA1c), low-density lipoprotein cholesterol (LDL-C), total cholesterol, smoking, and regular exercise. Model fit indices were acceptable (GFI=0.93, NFI=0.95, RMSEA=0.07).

Similar finding were observed for cDBP, with a direct effect of −0.088, an indirect effect of −0.004, and a total effect of −0.084 (Table S2). Model fit indices were also adequate (GFI = 0.91, NFI = 0.92, RMSEA = 0.06; Table S4; Figure 3). These results indicate consistent pathways for both cSBP and cDBP, with copper partially mediating the inverse association between 25(OH)D and central BP.

**Figure 3.**
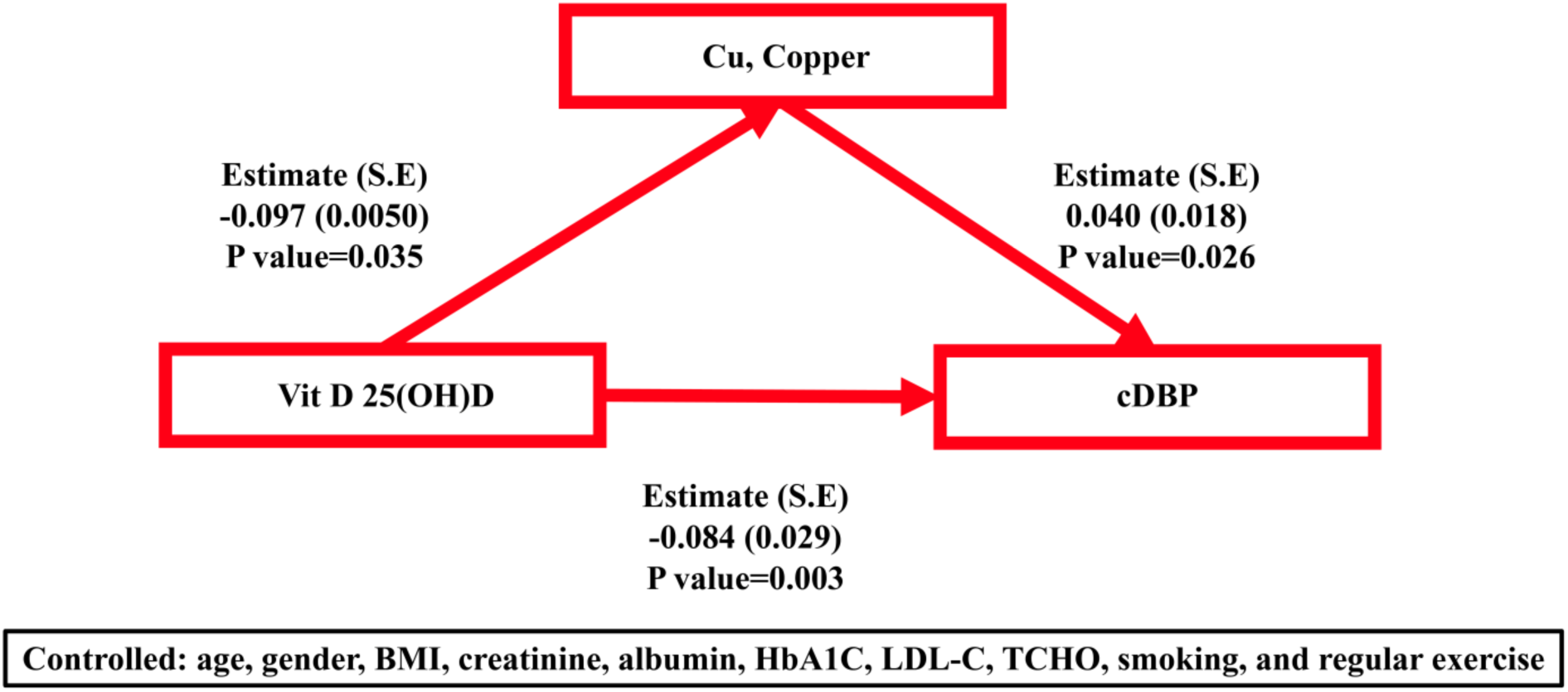
Structural equation modeling (SEM) illustrating the association amongc serum 25-hydroxyvitamin D [25(OH)D], urinary copper (Cu), and central diastolic blood pressure (cDBP). Arrows indicate direct and indirect pathways with corresponding standardized coefficients (standard errors) and P values. The model was adjusted for age, sex, body mass index, serum creatinine, albumin, glycated hemoglobin (HbA1c), low-density lipoprotein cholesterol (LDL-C), total cholesterol, smoking, and regular exercise. Model fit indices were acceptable (GFI=0.91, NFI=0.92, RMSEA=0.06).

## Discussion

This study yielded four principal findings. First, more than half (54.1%) of healthy young adults in Taiwan had vitamin D deficiency. Second, lower 25(OH)D levels were significantly associated with higher central systolic, diastolic, and mean arterial pressures, particularly among women and participants with BMI < 24 kg/m^2^. Third, a U-shaped relationship was observed between 25(OH)D and hypertension risk, with the lowest risk around 20.1 ng/mL. Finally, copper partially mediated the inverse association between vitamin D deficiency and central blood pressure.

The prevalence of vitamin D deficiency in our cohort was substantially higher than that previously reported in Taiwan (22% in the general population and 38.4% among adults aged 30–39 years)^24^ and was comparable to data observed in the United States^40^, highlighting vitamin D deficiency as an important public health concern.

Mechanistically, vitamin D deficiency may elevate blood pressure through activation of the renin–angiotensin–aldosterone system, secondary hyperparathyroidism, and impaired vascular smooth muscle relacation.^41,42^ Prior observational studies and meta-analyses have reported inverse associations between vitamin D and blood pressure, although large supplementation trials in the general population^12,13,43,44^ have not consistently demonstrated reductions in blood pressure.^45^ Nonetheless, longer-term supplementation among deficient individuals may lower central blood pressure.^46^ The U-shaped associations observed in this study parallel finding linking vitamin D with diabetes, cardiovascular disease, and mortality, suggesting that both deficiency and excess may be harmful.^47^ Importantly, we measures central rather than brachial blood pressure. Central pressure provides a more accurate reflection of target-organ load and cardiovascular risk than brachial pressure. In young adults, it may differ considerably from brachial pressure because of greater vascular elasticity and pulse wave amplification. Our finding of a U-shaped association with hypertension risk, with a nadir at 20.1 ng/mL, are consistent with prior U.S. data (<17.8 ng/mL) and NHANES III analyses showing a reverse J-shaped relationship with mortality.^48^ These results suggest that maintaining vitamin D at moderate levels may be optimal for cardiovascular health.^49^

Although subgroup patterns suggested stronger associations in men, formal interaction analyses showed no significant effect modification by age or gender. Interestingly, the inverse relationship between 25(OH)D and cBP appeared more pronounced in women, highlighting potential differences between central hemodynamics and clinical hypertension as outcomes. Taken together, our findings indicated that vitamin D status is linked to blood pressure regulation across demographic groups, but supplementation strategies may not need to be tailored solely on the basis of age or gender.

We further examined trace elements and found that among eight heavy metals measured, only urinary copper was significantly associated with both 25(OH)D and central blood pressure. Copper homeostasis is critical for vascular physiology, and excess copper may promote oxidative stress, endothelial dysfunction, and increased vascular tone. Our mediation analysis demonstrated that copper partially mediated the relationship between 25(OH)D and central blood pressure, although the indirect effect was modest. This observation aligns with prior evidence from Arab adults reporting an inverse relationship between vitamin D and copper levels,^50^ and supports the hypothesis the trace elements may play a role in vitamin D-related vascular regulation.

This study has several limitations. First, its cross-sectional design preclude causal inference. Second, the modest sample size may limit generalizability, and replication in larger cohorts is warranted. Third, the absence of longitudinal data restricts the ability to assess temporal changes. Finally, the relatively small absolute differences in blood pressure likely reflect the generally healthy status of the study population.

## Conclusion

In this cohort of healthy young adults in Taiwan, vitamin D deficiency was independently associated with higher central blood pressure. A non-linear, U-shaped association was observed between 25(OH)D and hypertension risk, with the lowest risk at 20.1 ng/mL. Structural equation modeling further suggested that copper may partially mediate this relationship. These findings highlight the importance of maintaining optimal vitamin D levels for blood pressure regulation, with copper acting as a potential mediator in this association.

## Data Availability

The data that support the findings of this study are available from the corresponding author upon reasonable request. Individual-level participant data are not publicly available due to privacy and ethical restrictions imposed by the institutional review board of National Taiwan University Hospital.

## Reference

1. Ezzati M, Lopez AD, Rodgers A, Vander Hoorn S, Murray CJ. Selected major risk factors and global and regional burden of disease. Lancet. 2002;360:1347–1360. doi: 10.1016/s0140-6736(02)11403-6

2. Mancia G, Kreutz R, Brunström M, Burnier M, Grassi G, Januszewicz A, Muiesan ML, Tsioufis K, Agabiti-Rosei E, Algharably EAE, et al. 2023 ESH Guidelines for the management of arterial hypertension The Task Force for the management of arterial hypertension of the European Society of Hypertension: Endorsed by the International Society of Hypertension (ISH) and the European Renal Association (ERA). J Hypertens. 2023;41:1874–2071. doi: 10.1097/hjh.0000000000003480

3. Terentes-Printzios D, Gardikioti V, Vlachopoulos C. Central Over Peripheral Blood Pressure: An Emerging Issue in Hypertension Research. Heart Lung Circ. 2021;30:1667–1674. doi: 10.1016/j.hlc.2021.07.019

4. Andress DL. Vitamin D in chronic kidney disease: a systemic role for selective vitamin D receptor activation. Kidney Int. 2006;69:33–43. doi: 10.1038/sj.ki.5000045

5. Ross AC, Manson JE, Abrams SA, Aloia JF, Brannon PM, Clinton SK, Durazo-Arvizu RA, Gallagher JC, Gallo RL, Jones G, et al. The 2011 report on dietary reference intakes for calcium and vitamin D from the Institute of Medicine: what clinicians need to know. J Clin Endocrinol Metab. 2011;96:53–58. doi: 10.1210/jc.2010-2704

6. Holick MF, Binkley NC, Bischoff-Ferrari HA, Gordon CM, Hanley DA, Heaney RP, Murad MH, Weaver CM. Evaluation, treatment, and prevention of vitamin D deficiency: an Endocrine Society clinical practice guideline. J Clin Endocrinol Metab. 2011;96:1911–1930. doi: 10.1210/jc.2011-0385

7. Norman PE, Powell JT. Vitamin D and cardiovascular disease. Circ Res. 2014;114:379–393. doi: 10.1161/circresaha.113.301241

8. Bouillon R, Carmeliet G, Verlinden L, van Etten E, Verstuyf A, Luderer HF, Lieben L, Mathieu C, Demay M. Vitamin D and human health: lessons from vitamin D receptor null mice. Endocr Rev. 2008;29:726–776. doi: 10.1210/er.2008-0004

9. Xiang W, Kong J, Chen S, Cao LP, Qiao G, Zheng W, Liu W, Li X, Gardner DG, Li YC. Cardiac hypertrophy in vitamin D receptor knockout mice: role of the systemic and cardiac renin-angiotensin systems. Am J Physiol Endocrinol Metab. 2005;288:E125–132. doi: 10.1152/ajpendo.00224.2004

10. Forman JP, Williams JS, Fisher ND. Plasma 25-hydroxyvitamin D and regulation of the renin-angiotensin system in humans. Hypertension. 2010;55:1283–1288. doi: 10.1161/hypertensionaha.109.148619

11. McMullan CJ, Borgi L, Curhan GC, Fisher N, Forman JP. The effect of vitamin D on renin-angiotensin system activation and blood pressure: a randomized control trial. J Hypertens. 2017;35:822–829. doi: 10.1097/hjh.0000000000001220

12. Judd SE, Nanes MS, Ziegler TR, Wilson PW, Tangpricha V. Optimal vitamin D status attenuates the age-associated increase in systolic blood pressure in white Americans: results from the third National Health and Nutrition Examination Survey. Am J Clin Nutr. 2008;87:136–141. doi: 10.1093/ajcn/87.1.136

13. Che J, Tong J, Kuang X, Zheng C, Zhou R, Song J, Zhan X, Liu Z. Relationship between serum 25-hydroxyvitamin D concentrations and blood pressure among US adults without a previous diagnosis of hypertension: evidence from NHANES 2005-2018. Front Nutr. 2023;10:1265662. doi: 10.3389/fnut.2023.1265662

14. Scragg R, Sowers M, Bell C. Serum 25-hydroxyvitamin D, ethnicity, and blood pressure in the Third National Health and Nutrition Examination Survey. Am J Hypertens. 2007;20:713–719. doi: 10.1016/j.amjhyper.2007.01.017

15. Hintzpeter B, Mensink GB, Thierfelder W, Müller MJ, Scheidt-Nave C. Vitamin D status and health correlates among German adults. Eur J Clin Nutr. 2008;62:1079–1089. doi: 10.1038/sj.ejcn.1602825

16. Moore CE, Liu Y. Elevated systolic blood pressure of children in the United States is associated with low serum 25-hydroxyvitamin D concentrations related to body mass index: National Health and Examination Survey 2007-2010. Nutr Res. 2017;38:64–70. doi: 10.1016/j.nutres.2017.01.008

17. Vishnu A, Ahuja V. Vitamin D and Blood Pressure Among U.S. Adults: A Cross-sectional Examination by Race/Ethnicity and Gender. Am J Prev Med. 2017;53:670–679. doi: 10.1016/j.amepre.2017.07.006

18. Karadeniz Y, Özpamuk-Karadeniz F, Ahbab S, Ataoğlu E, Can G. Vitamin D Deficiency Is a Potential Risk for Blood Pressure Elevation and the Development of Hypertension. Medicina (Kaunas). 2021;57. doi: 10.3390/medicina57121297

19. Chen J, Jiang Y, Shi H, Peng Y, Fan X, Li C. The molecular mechanisms of copper metabolism and its roles in human diseases. Pflugers Arch. 2020;472:1415–1429. doi: 10.1007/s00424-020-02412-2

20. Fukai T, Ushio-Fukai M, Kaplan JH. Copper transporters and copper chaperones: roles in cardiovascular physiology and disease. Am J Physiol Cell Physiol. 2018;315:C186–c201. doi: 10.1152/ajpcell.00132.2018

21. He P, Li H, Liu C, Liu M, Zhang Z, Zhang Y, Zhou C, Li Q, Ye Z, Wu Q, et al. U-shaped association between dietary copper intake and new-onset hypertension. Clin Nutr. 2022;41:536–542. doi: 10.1016/j.clnu.2021.12.037

22. Miao Q, Zhang J, Yun Y, Wu W, Luo C. Association between copper intake and essential hypertension: dual evidence from Mendelian randomization analysis and the NHANES database. Front Nutr. 2024;11:1454669. doi: 10.3389/fnut.2024.1454669

23. Chen CW, Shu CC, Han YY, Hsu SH, Hwang JS, Su TC. Mediated relationship between Vitamin D deficiency and reduced pulmonary function by copper in Taiwanese young adults. Ecotoxicol Environ Saf. 2024;285:117034. doi: 10.1016/j.ecoenv.2024.117034

24. Lee MJ, Hsu HJ, Wu IW, Sun CY, Ting MK, Lee CC. Vitamin D deficiency in northern Taiwan: a community-based cohort study. BMC Public Health. 2019;19:337. doi: 10.1186/s12889-019-6657-9

25. Chien KL, Hsu HC, Chen PC, Lin HJ, Su TC, Chen MF, Lee YT. Total 25-hydroxyvitamin D concentration as a predictor for all-cause death and cardiovascular event risk among ethnic Chinese adults: a cohort study in a Taiwan community. PLoS One. 2015;10:e0123097. doi: 10.1371/journal.pone.0123097

26. Wei JN, Sung FC, Lin CC, Lin RS, Chiang CC, Chuang LM. National surveillance for type 2 diabetes mellitus in Taiwanese children. JAMA. 2003;290:1345–1350. doi: 10.1001/jama.290.10.1345

27. Lin CY, Wang CK, Sung FC, Su TC. The Association among Urinary Lead and Cadmium, Serum Adiponectin, and Serum Apoptotic Microparticles in a Young Taiwanese Population. Nutrients. 2023;15. doi: 10.3390/nu15214528

28. Lin CY, Lee HL, Wang C, Sung FC, Su TC. Examining the impact of polyfluoroalkyl substance exposure on erythrocyte profiles and its related nutrients: Insights from a prospective study on young Taiwanese. Environ Pollut. 2024;359:124576. doi: 10.1016/j.envpol.2024.124576

29. Tsao TM, Tsai MJ, Hwang JS, Su TC. Health effects of seasonal variation in cardiovascular hemodynamics among workers in forest environments. Hypertens Res. 2019;42:223–232. doi: 10.1038/s41440-018-0136-z

30. Brinton TJ, Cotter B, Kailasam MT, Brown DL, Chio SS, O’Connor DT, DeMaria AN. Development and validation of a noninvasive method to determine arterial pressure and vascular compliance. Am J Cardiol. 1997;80:323–330. doi: 10.1016/s0002-9149(97)00353-6

31. Brinton TJ, Walls ED, Chio SS. Validation of pulse dynamic blood pressure measurement by auscultation. Blood Press Monit. 1998;3:121–124.

32. Marcus RH, Korcarz C, McCray G, Neumann A, Murphy M, Borow K, Weinert L, Bednarz J, Gretler DD, Spencer KT, et al. Noninvasive method for determination of arterial compliance using Doppler echocardiography and subclavian pulse tracings. Validation and clinical application of a physiological model of the circulation. Circulation. 1994;89:2688–2699. doi: 10.1161/01.cir.89.6.2688

33. Tsao TM, Hwang JS, Chen CY, Lin ST, Tsai MJ, Su TC. Urban climate and cardiovascular health: Focused on seasonal variation of urban temperature, relative humidity, and PM(2.5) air pollution. Ecotoxicol Environ Saf. 2023;263:115358. doi: 10.1016/j.ecoenv.2023.115358

34. Tsao TM, Hwang JS, Tsai MJ, Lin ST, Wu C, Su TC. Seasonal Effects of High-Altitude Forest Travel on Cardiovascular Function: An Overlooked Cardiovascular Risk of Forest Activity. Int J Environ Res Public Health. 2021;18. doi: 10.3390/ijerph18189472

35. Tsao TM, Hwang JS, Lin ST, Wu C, Tsai MJ, Su TC. Forest Bathing Is Better than Walking in Urban Park: Comparison of Cardiac and Vascular Function between Urban and Forest Parks. Int J Environ Res Public Health. 2022;19. doi: 10.3390/ijerph19063451

36. Wang TD, Chiang CE, Chao TH, Cheng HM, Wu YW, Wu YJ, Lin YH, Chen MY, Ueng KC, Chang WT, et al. 2022 Guidelines of the Taiwan Society of Cardiology and the Taiwan Hypertension Society for the Management of Hypertension. Acta Cardiol Sin. 2022;38:225–325. doi: 10.6515/acs.202205_38(3).20220321a

37. Writing Committee M, Jones DW, Ferdinand KC, Taler SJ, Johnson HM, Shimbo D, Abdalla M, Altieri MM, Bansal N, Bello NA, et al. 2025 AHA/ACC/AANP/AAPA/ABC/ACCP/ACPM/AGS/AMA/ASPC/NMA/PCNA/SGIM Guideline for the Prevention, Detection, Evaluation and Management of High Blood Pressure in Adults: A Report of the American College of Cardiology/American Heart Association Joint Committee on Clinical Practice Guidelines. Hypertension. 2025;82:e212–e316. doi: 10.1161/HYP.0000000000000249

38. Tsugawa N, Suhara Y, Kamao M, Okano T. Determination of 25-hydroxyvitamin D in human plasma using high-performance liquid chromatography--tandem mass spectrometry. Anal Chem. 2005;77:3001–3007. doi: 10.1021/ac048249c

39. Wise SA, Camara JE, Sempos CT, Lukas P, Le Goff C, Peeters S, Burdette CQ, Nalin F, Hahm G, Durazo-Arvizu RA, et al. Vitamin D Standardization Program (VDSP) intralaboratory study for the assessment of 25-hydroxyvitamin D assay variability and bias. The Journal of Steroid Biochemistry and Molecular Biology. 2021;212:105917. doi: 10.1016/j.jsbmb.2021.105917

40. Nair R, Maseeh A. Vitamin D: The “sunshine” vitamin. J Pharmacol Pharmacother. 2012;3:118–126. doi: 10.4103/0976-500x.95506

41. Snijder MB, Lips P, Seidell JC, Visser M, Deeg DJ, Dekker JM, van Dam RM. Vitamin D status and parathyroid hormone levels in relation to blood pressure: a population-based study in older men and women. J Intern Med. 2007;261:558–565. doi: 10.1111/j.1365-2796.2007.01778.x

42. Wong MS, Delansorne R, Man RY, Vanhoutte PM. Vitamin D derivatives acutely reduce endothelium-dependent contractions in the aorta of the spontaneously hypertensive rat. Am J Physiol Heart Circ Physiol. 2008;295:H289–296. doi: 10.1152/ajpheart.00116.2008

43. He JL, Scragg RK. Vitamin D, parathyroid hormone, and blood pressure in the National Health and Nutrition Examination Surveys. Am J Hypertens. 2011;24:911–917. doi: 10.1038/ajh.2011.73

44. Kunutsor SK, Apekey TA, Steur M. Vitamin D and risk of future hypertension: meta-analysis of 283,537 participants. Eur J Epidemiol. 2013;28:205–221. doi: 10.1007/s10654-013-9790-2

45. Scragg R, Slow S, Stewart AW, Jennings LC, Chambers ST, Priest PC, Florkowski CM, Camargo CA, Jr., Murdoch DR. Long-term high-dose vitamin D3 supplementation and blood pressure in healthy adults: a randomized controlled trial. Hypertension. 2014;64:725–730. doi: 10.1161/hypertensionaha.114.03466

46. Sluyter JD, Camargo CA, Jr., Stewart AW, Waayer D, Lawes CMM, Toop L, Khaw KT, Thom SAM, Hametner B, Wassertheurer S, et al. Effect of Monthly, High-Dose, Long-Term Vitamin D Supplementation on Central Blood Pressure Parameters: A Randomized Controlled Trial Substudy. J Am Heart Assoc. 2017;6. doi: 10.1161/jaha.117.006802

47. Sofianopoulou EK, Stephen K; Afzal, Shoaib; Jiang, Tao; Gill, Dipender; Gundersen, Thomas E; …; Butterworth Adam S; Burgess, Stephen. Estimating dose-response relationships for vitamin D with coronary heart disease, stroke, and all-cause mortality: observational and Mendelian randomisation analyses. Lancet Diabetes Endocrinol. 2024;12:e2–e11. doi: 10.1016/s2213-8587(23)00287-5

48. Melamed ML, Michos ED, Post W, Astor B. 25-hydroxyvitamin D levels and the risk of mortality in the general population. Arch Intern Med. 2008;168:1629–1637. doi: 10.1001/archinte.168.15.1629

49. Sempos CT, Durazo-Arvizu RA, Dawson-Hughes B, Yetley EA, Looker AC, Schleicher RL, Cao G, Burt V, Kramer H, Bailey RL, et al. Is there a reverse J-shaped association between 25-hydroxyvitamin D and all-cause mortality? Results from the U.S. nationally representative NHANES. J Clin Endocrinol Metab. 2013;98:3001–3009. doi: 10.1210/jc.2013-1333

50. Al-Daghri NM, Alfadul H, Kattak MNK, Yakout S. Vitamin D and its influence in circulating trace minerals among Arab adults with or without adequate vitamin D levels. Journal of King Saud University - Science. 2022;34:102012. doi: 10.1016/j.jksus.2022.102012

